# Diagnostic Value Of Urine Microscopy Leukocyte Findings in Predicting Urine Culture Positivity in Routine Clinical Practice

**DOI:** 10.64898/2026.02.01.26344693

**Authors:** Sadık Portakal, Basri Cakiroglu, Ramazan Gozukucuk, Selami Aydın

## Abstract

**Objective:** Urine cultures are frequently requested at an early stage in primary care and outpatient settings, often without a comprehensive clinical assessment. This practice increases healthcare costs and laboratory workload and may lead to misleading results due to asymptomatic bacteriuria and specimen contamination.

This study aimed to evaluate whether routinely reported microscopic urinary leukocyte findings can predict urine culture positivity under real-world clinical conditions. The distribution of isolated microorganisms and the frequency of mixed or contaminated growth were assessed.

**Methods:** This retrospective, laboratory-based diagnostic accuracy study included all urine samples sent for culture over a one-year period at a tertiary care hospital, provided concurrent microscopic urinalysis was available. No additional clinical exclusion criteria were applied to reflect the routine practice. Leukocyte findings were reported semi-quantitatively and analyzed both categorically and as approximate numerical values. The urine culture results were classified as positive, negative, or mixed/contaminated growth. The diagnostic performance was evaluated using receiver operating characteristic (ROC) curve analysis.

**Results:** A total of 8,478 urine samples were analyzed in this study. Urine culture positivity was detected in 2,666 (31.4%) samples, whereas 5, 812 (68.6%) showed no growth. Culture positivity increased significantly with higher leukocyte levels (p < 0.001), ranging from 13.1% in the lowest category to 83.1% in samples with abundant leukocytes. ROC analysis demonstrated an acceptable discriminative performance (AUC = 0.747). The Youden index identified an optimal threshold of approximately 5.5 leukocytes per high-power field, with a sensitivity of 60.4% and a specificity of 77.8%. Mixed or contaminated growth was the most common finding among culture-positive samples (43.5%), followed by *Escherichia coli* (29.5%).

**Conclusion:** Microscopic urinary leukocyte findings were significantly associated with urine culture positivity and demonstrated acceptable predictive performance in real-world clinical practice settings. Although leukocyte microscopy alone is not diagnostic, it may support more selective urine culture ordering, reduce contamination, and contribute to rational diagnosis and antimicrobial management in primary care.

## Introduction

Urinary tract infections (UTIs) are among the most common reasons for visits to primary care and outpatient clinics and account for a substantial proportion of antibiotic prescriptions worldwide (1,2). In routine clinical practice, urine cultures are frequently requested during the initial evaluation, often before a detailed clinical assessment is performed. Although urine culture remains the reference standard for pathogen identification, indiscriminate ordering may increase laboratory workload and healthcare costs, delay clinical decision-making, and contribute to misinterpretation of mixed or contaminated specimens (3–5).

In primary care and outpatient settings, clinicians often need to make diagnostic and treatment decisions using limited clinical and laboratory information. Complete urinalysis and urine microscopy are widely used because they are inexpensive, rapidly available, and easily accessible in daily practice (6). Among microscopic findings, urinary leukocytes (pyuria) are commonly interpreted as markers of urinary tract inflammation and potential infection. However, the clinical utility of pyuria for predicting bacteriuria remains debated, particularly in patients with non-specific symptoms or discordant clinical findings (7,8,20). Accordingly, previous studies have reported highly variable diagnostic performance for pyuria, with sensitivity and specificity estimates differing by study design, patient population, and clinical context (9–11).

Most available evidence derives from selected populations such as hospitalized patients, pediatric cohorts, or patients with clearly defined symptomatic UTIs. In contrast, real-world outpatient practice includes a broader spectrum of indications for ordering urine cultures—such as screening, follow-up testing, or vague urinary complaints—where evidence is comparatively limited (12). From a primary care perspective, identifying patients with a low probability of culture positivity is particularly important. Overuse of urine cultures increases the detection of asymptomatic bacteriuria and mixed/contaminated growth, which may prompt unnecessary antibiotic prescribing and contribute to antimicrobial resistance (13,14).

Despite the widespread use of urine microscopy, no universally accepted leukocyte threshold reliably predicts urine culture positivity in routine practice (15,16,19). Moreover, mixed or contaminated growth patterns—often related to suboptimal sampling—create diagnostic uncertainty and may lead to inappropriate management decisions (17). Therefore, the present study aimed to assess the predictive value of microscopic urinary leukocyte findings for urine culture positivity using real-world laboratory data collected over one year. Secondary objectives were to evaluate diagnostic performance using receiver operating characteristic (ROC) analysis and to describe microorganism distribution, including mixed/contaminated growth patterns, among culture-positive samples.

among culture-positive samples.

## Methods and Analysis

### Study design and setting

This retrospective study was conducted at Hisar Intercontinental Hospital and included urine samples from patients presenting to the urology, family medicine, internal medicine, and infectious diseases outpatient clinics over a one-year period. The study was designed as a laboratory based diagnostic accuracy analysis to evaluate the diagnostic value of microscopic urine leukocyte levels for predicting urine culture positivity under routine clinical conditions.

### Data source and study population

Routinely collected laboratory data were retrieved from the hospital laboratory information system. All urine samples submitted for **urine culture** with **concurrent microscopic urinalysis** during the study period were screened. Each urine sample was analyzed as an independent observation to reflect routine practice.

### Inclusion and exclusion criteria

**Inclusion criteria** were:

1. urine samples obtained from outpatient clinic attendees (urology, internal medicine, infectious diseases) during the study period;
2. submission for urine culture with a concurrent microscopic urinalysis performed as part of routine care;
3. complete culture reporting and documented microscopic leukocyte results.

**Exclusion criteria** were limited to:

1. missing or incomplete laboratory data (absent culture result or missing leukocyte reporting);
2. rejected or uninterpretable specimens due to pre-analytical/analytical issues documented by the laboratory (e.g., insufficient volume, leakage, labeling/identification discrepancies, or processing failure), when applicable;
3. duplicate laboratory records originating from the same accession number; in such cases, only the final validated report was retained.

No additional clinical exclusions (e.g., age, sex, antibiotic exposure, comorbidities) were applied to preserve real-world representativeness.

### Laboratory procedures

Microscopic urinalysis and urine culture were performed as part of the routine workflow of the hospital laboratory. Urine samples were processed according to standard operating procedures in place during the study period. Microscopic examination was conducted using (manual light microscopy / an automated urine analyzer with microscopic confirmation), and leukocytes were reported semi-quantitatively as rare, 1–2, 3–4, 5–6, ≥10 leukocytes per high-power field (HPF), or “abundant,” as documented in the laboratory information system. For analysis, leukocyte findings were treated as ordinal categorical variables; for ROC analyses, approximate numerical values were assigned to each category based on predefined ranges.

Urine culture was performed using routine microbiological methods. Samples were inoculated on appropriate culture media and incubated under standard conditions. Culture reports were classified as positive, negative (no growth), or mixed/contaminated growth according to the laboratory reporting format. When positive, microorganisms were identified using routine identification methods (e.g., conventional biochemical testing and/or automated identification systems such as VITEK/MALDI-TOF, if available), and results were recorded at the species/genus level as provided in the reports. Mixed/contaminated growth patterns were retained as a separate category, reflecting real-world reporting.

### Blinding

The microscopic urinalysis (index test) and urine culture (reference standard) were performed and reported independently as part of the routine laboratory workflow. Laboratory personnel interpreting urine microscopy were not involved in urine culture processing or reporting, and culture results were not available at the time of microscopic assessment. Likewise, urine culture interpretation was performed without access to microscopic leukocyte results.

### Outcomes

The **primary outcome** was urine culture positivity. The primary predictor was the microscopic urine leukocyte level. Secondary outcomes included (i) diagnostic performance measures of leukocyte levels at different thresholds and (ii) distribution of microorganisms among culture-positive samples, including the frequency of mixed/contaminated growth.

### Statistical analysis

Continuous variables were assessed for distribution using histograms/Q–Q plots and the Shapiro–Wilk test. Normally distributed variables were summarized as mean ± SD; non-normally distributed variables as median (IQR). Categorical variables were presented as counts and percentages.

Associations between leukocyte categories and culture results were evaluated using the chi-square test (or Fisher’s exact test when expected cell counts were <5). For comparisons of non-normally distributed continuous variables between two groups, the Mann–Whitney U test was used. A trend across ordered leukocyte categories in relation to culture positivity was evaluated using a chi-square test for trend, where applicable. All tests were two-sided, and statistical significance was set at *P* < 0.05.

Diagnostic accuracy of microscopic leukocyte levels for predicting culture positivity was evaluated using receiver operating characteristic (ROC) curve analysis. Discrimination was quantified using the area under the ROC curve (AUC) with 95% confidence intervals (CIs). The optimal cutoff value was determined using the Youden index (J = sensitivity + specificity − 1). At the selected cutoff, sensitivity, specificity, positive predictive value (PPV), negative predictive value (NPV), positive likelihood ratio (LR+), and negative likelihood ratio (LR−) were calculated with 95% CIs. The primary ROC comparison was performed between culture-positive and culture-negative samples; mixed/contaminated growth was analyzed descriptively as a separate category.

Missing data were handled by complete-case analysis, as samples with incomplete microscopy or culture data were excluded by design. This study was reported in accordance with the STARD 2015 (Standards for Reporting Diagnostic Accuracy Studies) reporting guideline. The STARD reporting checklist was used during manuscript preparation and editing and is provided as a supplementary file.(23)

### Ethics

This study was conducted in accordance with the Declaration of Helsinki and approved by the Ethics Committee of Hisar Intercontinental Hospital on October 15, 2025 (Approval No: 25-46). Given the retrospective design and use of anonymized routine laboratory data, the requirement for informed consent was waived by the Ethics Committee.

### Trial registration

Not applicable.

This study was a retrospective observational analysis using existing routinely collected laboratory data and did not involve prospective assignment of participants to any intervention. Therefore, in accordance with the ICMJE definition of a clinical trial, trial registration was not required.

## Results

During the one-year study period, a total of 8,478 urine samples with complete microscopy and culture results were included in the analysis. Baseline demographic and clinical characteristics of the study population are summarized in Table 1. Age distribution parameters were calculated from the complete dataset and extrapolated to the overall cohort to reflect a real-world outpatient population.

**Table 1.** Baseline characteristics of the study population.

| Characteristic | Value |
| --- | --- |
| **Total urine samples. n** | 8,478 |
| **Sex. n (%)** |  |
| Female | 6,076 (71.7%) |
| Male | 2,402 (28.3%) |
| **Age. years** |  |
| Mean ± SD | 36.9 ± 24.4 |
| Median (IQR) | 33 (20–56) |
| **Care setting** | Outpatient |
| **Care level** | Tertiary care |
| **Study design** | Retrospective, laboratory-based |
| **Center type** | Single-center |

Overall, 2,666 samples (31.4%) yielded positive urine culture results, while 5,812 samples (68.6%) showed no bacterial growth. Urine culture positivity demonstrated a clear stepwise increase with rising microscopic leukocyte counts (Table 2). Culture positivity rates were lowest in samples with rare or low leukocyte levels and increased progressively across higher leukocyte categories, reaching a maximum positivity rate of 83.1% in samples with abundant leukocytes. The association between microscopic leukocyte levels and urine culture positivity was statistically significant (χ^2^ test, p < 0.001).

**Table 2.** Distribution of urine culture results according to microscopic leukocyte findings.

| Leukocyte category | Total (n) | Culture positive n (%) | Culture negative n (%) |
| --- | --- | --- | --- |
| Nadir | 411 | 54 (13.1%) | 357 (86.9%) |
| 1–2 / HPF | 2,019 | 294 (14.6%) | 1,725 (85.4%) |
| 2–3 / HPF | 1,664 | 326 (19.6%) | 1,338 (80.4%) |
| 3–4 / HPF | 862 | 214 (24.8%) | 648 (75.2%) |
| 4–5 / HPF | 489 | 137 (28.0%) | 352 (72.0%) |
| 5–6 / HPF | 393 | 130 (33.1%) | 263 (66.9%) |
| 8–10 / HPF | 351 | 124 (35.3%) | 227 (64.7%) |
| 10–15 / HPF | 320 | 148 (46.3%) | 172 (53.7%) |
| 15–20 / HPF | 231 | 122 (52.8%) | 109 (47.2%) |
| 20–25 / HPF | 109 | 72 (66.1%) | 37 (33.9%) |
| 25–30 / HPF | 84 | 53 (63.1%) | 31 (36.9%) |
| Abundant leukocytes | 852 | 708 (83.1%) | 144 (16.9%) |
| **Total** | **8,478** | **2,666 (31.4%)** | **5,812 (68.6%)** |

The rate of urine culture positivity increased progressively with higher microscopic leukocyte counts ( $\chi^2$ , $p < 0.001$ ). Samples with abundant leukocytes had the highest culture positivity rate (83.1%).

Analysis of culture-positive samples revealed that mixed or contaminated growth patterns constituted the largest proportion of samples (43.5%). Among samples with a single identified pathogen, Escherichia coli was the most frequently isolated microorganism (29.5% of samples). Other pathogens, including Klebsiella spp., Enterococcus spp., Proteus spp., Candida spp., and Pseudomonas aeruginosa, were detected at lower frequencies (Table 3). Abundant growth was observed in most isolates, supporting the presence of clinically significant infections. Notably, the detection of Candida species and non-fermentative gram-negative bacteria was consistent with hospitalized and clinically complicated patient populations.

**Table 3.** Distribution of microorganisms in culture-positive urine samples.

| Microorganism | n | % |
| --- | --- | --- |
| Mixed / contaminated growth | 1,156 | 43.5 |
| Escherichia coli | 782 | 29.5 |
| Klebsiella spp. | 128 | 4.8 |
| Streptococcus agalactiae | 55 | 2.1 |
| Enterococcus spp. | 47 | 1.8 |
| Proteus spp. | 29 | 1.1 |
| Pseudomonas aeruginosa | 18 | 0.7 |
| Other single organisms (candida , gram (-) bac.) | 451 | 16.5 |
| Total | 2,666 | 100 |

Among the culture-positive samples, mixed or contaminated growth patterns constituted the largest proportion (43.5%), followed by Escherichia coli (29.5%).

Receiver operating characteristic (ROC) curve analysis demonstrated that microscopic urine leukocyte count had an acceptable discriminatory ability for differentiating culture-positive from culture-negative samples, with an area under the curve (AUC) of 0.747 (Figure 1). The optimal leukocyte cutoff value, determined using the Youden index, was approximately 5.5 leukocytes per high-power field, with a sensitivity of 60.4% and specificity of 77.8%.

**Figure 1.**
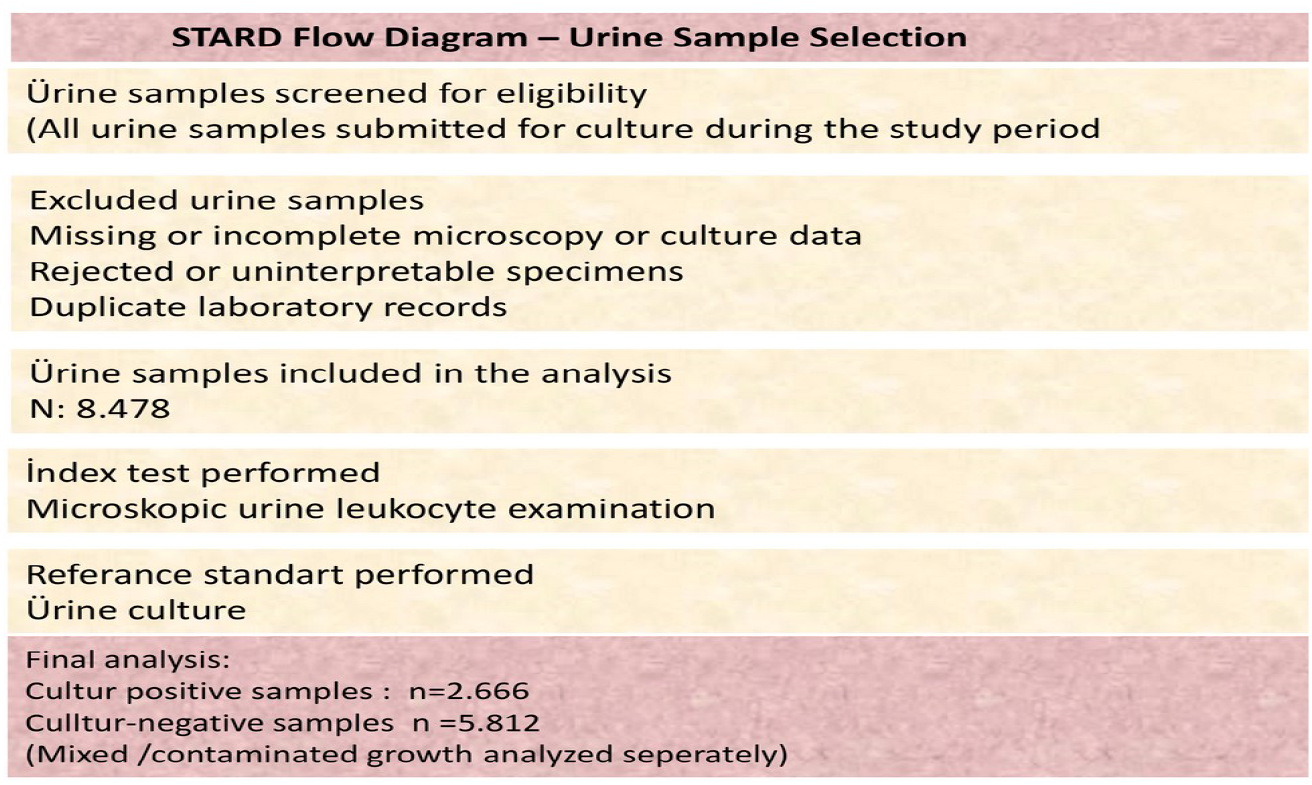
Flow diagram of urine sample selection and analysis.

**Figure 2.**
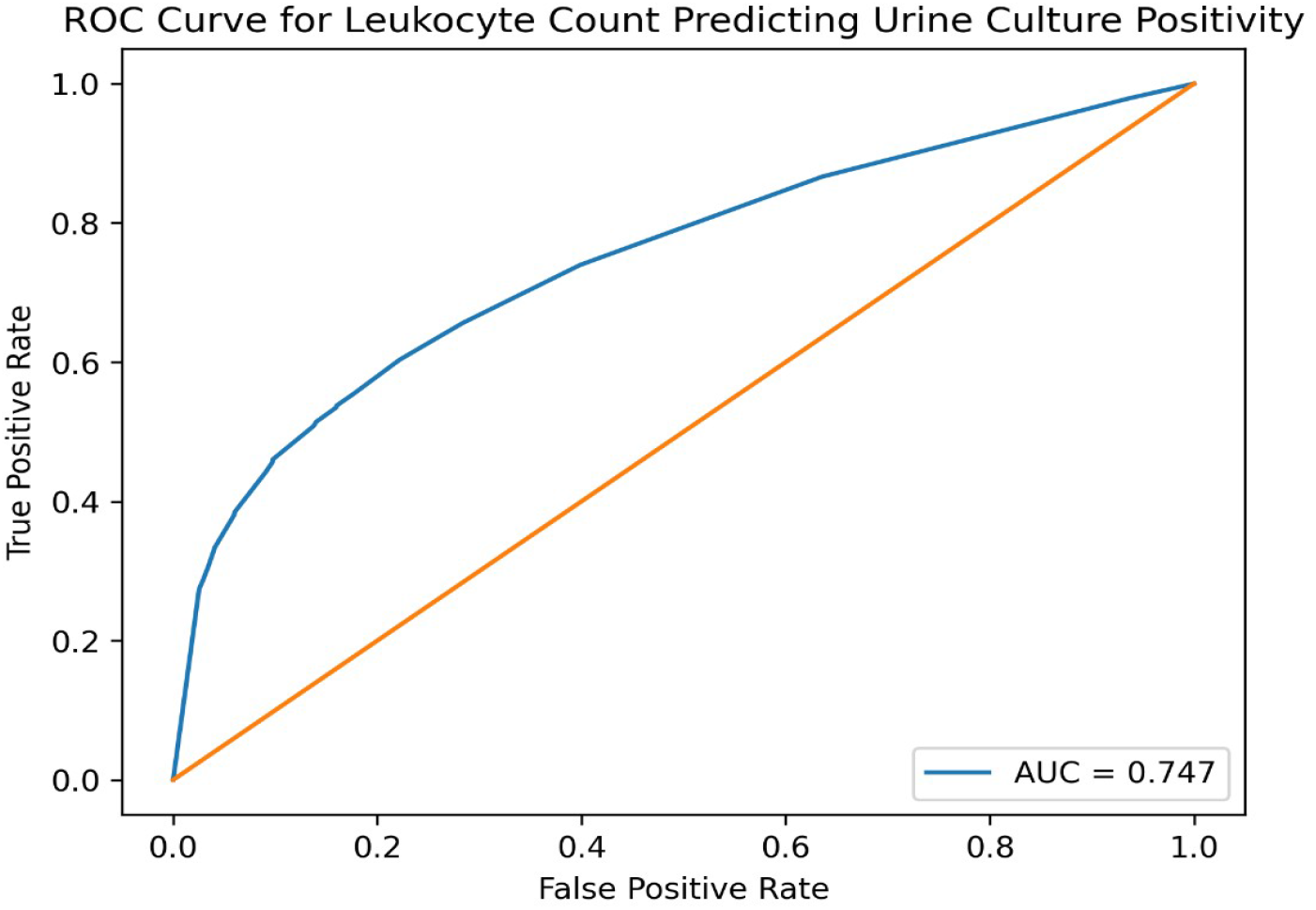
Receiver operating characteristic (ROC) curve of microscopic urine leukocyte count for predicting urine culture positivity.

This flow diagram illustrates the selection of urine samples included in the diagnostic accuracy analysis. All urine samples submitted for culture during the study period were screened. Samples with missing or incomplete microscopy or culture data, rejected or uninterpretable specimens, and duplicate laboratory records were excluded. A total of 8,478 urine samples with concurrent microscopic urinalysis and urine culture results were included in the final analysis.

Receiver operating characteristic (ROC) analysis showed that the microscopic leukocyte count was an acceptable predictor of urine culture positivity, with an area under the curve (AUC) of 0.747.

In the Youden index assessment, the optimal leukocyte threshold was approximately 5.5 leukocytes at high magnification, with a sensitivity of 60.4% and specificity of 77.8%. These findings suggest that higher leukocyte counts are associated with an increased probability of urine culture positivity; however, leukocyte counts alone are not sufficient for a definitive diagnosis.

## Discussion

In this study, we assessed the real-world diagnostic utility of routinely reported microscopic urinary leukocyte findings for predicting urine culture positivity in an outpatient setting. Using a large, unselected sample of urine cultures ordered in daily practice, we observed a strong graded relationship between leukocyte level and culture positivity. The stepwise rise in positivity across leukocyte categories supports urine microscopy as a clinically meaningful indicator of increasing likelihood of bacteriuria.

Urine cultures are commonly ordered early in outpatient care, sometimes before a complete clinical assessment is performed. While culture remains the reference standard for pathogen identification, indiscriminate ordering can increase laboratory workload and costs and can amplify the impact of mixed/contaminated specimens on clinical decision-making (3–5,18). From a diagnostic stewardship perspective, simple and rapidly available parameters that help identify low-probability samples are particularly valuable because they may reduce unnecessary cultures and downstream inappropriate antibiotic exposure (13,14).

Our ROC analysis demonstrated acceptable discrimination between culture-positive and culture-negative samples (AUC 0.747). The ROC-derived cutoff of approximately 5–6 leukocytes/HPF provides a pragmatic threshold range that may support selective culture ordering strategies in routine practice. Importantly, pyuria should not be considered diagnostic on its own; rather, leukocyte microscopy can be interpreted as supportive evidence that should be integrated with symptoms, physical examination, and clinical risk factors (6,20). This interpretation aligns with contemporary approaches advocating integration of clinical and laboratory information to optimize test utilization and antimicrobial stewardship (18–20).

A key observation in our dataset was the high proportion of mixed or contaminated growth among reports categorized as culture-positive. Similar findings have been attributed to suboptimal sampling technique and pre-analytical factors, which remain frequent challenges in outpatient care (17,21). Mixed growth patterns create diagnostic uncertainty and may lead to unnecessary repeat testing or empiric antibiotic prescribing. Incorporating leukocyte microscopy into pre-culture decision-making—particularly when leukocyte levels are low—may help reduce low-yield cultures and potentially decrease the burden of misleading mixed/contaminated results. In addition, these findings emphasize the need for improved patient instruction and standardized sampling protocols to reduce contamination.

Regarding microbiology, Escherichia coli was the most frequently isolated pathogen, consistent with established epidemiology of UTIs (22). The detection of Candida species and non-fermentative gram-negative bacteria suggests that the outpatient sample set likely included heterogeneous and potentially complicated clinical scenarios rather than exclusively uncomplicated community-acquired infections. This heterogeneity may enhance the practical relevance of the findings for routine practice, although it also reinforces the importance of clinical context when interpreting culture and microscopy results.

## Limitations

Several limitations should be considered when interpreting these results. First, due to the retrospective design and use of routine laboratory data, clinical variables (e.g., symptoms, comorbidities, pregnancy status, catheter use, and recent antibiotic exposure) were not available; therefore, the clinical significance of culture positivity in individual cases and potential confounding factors could not be assessed. Second, samples were analyzed as independent observations, and repeated samples from the same patient may have been included, which may violate independence assumptions. Third, leukocyte reporting was semi-quantitative, and assigning approximate numeric values for ROC analysis may introduce some measurement imprecision, although the consistent graded association across categories supports the robustness of the overall pattern. Finally, this single-center design may limit generalizability to other settings with different populations and laboratory workflows. Prospective multicenter studies incorporating clinical variables are needed to validate these findings and refine practical thresholds for diagnostic stewardship.

## Conclusion

Microscopic urine leukocyte levels showed a significant graded association with urine culture positivity in a large real-world outpatient dataset and demonstrated acceptable discrimination (AUC 0.747). While pyuria is not diagnostic alone, leukocyte microscopy—particularly around a threshold of >5–6 leukocytes/HPF may support more selective urine culture ordering when combined with clinical assessment. Prospective multicenter studies including symptom and risk-factor data are warranted to standardize practical thresholds and confirm clinical impact.

## Supporting information

https://docs.google.com/spreadsheets/d/1O_yIlRA7I_UcPZsiWbLuTEdcmeDIfPXR/edit?usp=drive_link&ouid=114451319444089279424&rtpof=true&sd=true

## Data Availability

The data used in this study are available from the corresponding author upon reasonable request.

https://docs.google.com/spreadsheets/d/1F4PhZzWikCFXUfVexRxd7BaJaTiLzU3f/edit?usp=sharing&ouid=114451319444089279424&rtpof=true&sd=true

## Declarations

### Ethics approval and consent to participate

Ethical approval for this study was obtained from the Ethics Committee of Hisar Intercontinental Hospital on **October 15, 2025 (Approval No: 25-46)**. Owing to the retrospective design of the study and the use of anonymized routine laboratory data, the requirement for informed consent was waived by the Ethics Committee.

### Consent for publication

Not applicable.

### Availability of data and materials

The datasets used and/or analyzed during the current study are available from the corresponding author upon reasonable request.

## Competing interests

The authors declare no conflict of interest.

## Funding

The authors received no financial support for the research, authorship, or publication of this article.

## Authors’ contributions

All authors contributed to the study’s conception and design. The authors performed data collection and analysis. The first draft of the manuscript was written by the authors, and all authors commented on previous versions of the manuscript. All authors have read and approved the final manuscript.

## Acknowledgements

I would like to express my sincere appreciation to my wife, Ferhan Portakal, and my daughter, Elif Gülderen Portakal, for their patience, understanding, and support during the preparation of this manuscript. I am also grateful to my parents for their continuous support. Furthermore, I would like to thank the Hospital Ethics Committee for their guidance and approval of this study. I also extend my thanks to Salahattin Öztekin, Head of the Hospital Information Technologies Department, for his valuable assistance during data collection.

## Notes

### Competing Interest Statement

The authors have declared no competing interest.

### Funding Statement

This study received no external funding.

### Author Declarations

Ethical approval for this study was obtained from the Ethics Committee of Hisar Intercontinental Hospital on October 15, 2025 (Approval No: 25-46). Owing to the retrospective design of the study and the use of anonymized routine laboratory data, the requirement for informed consent was waived by the Ethics Committee.

## References

1. Gözüküçük R, Çakıroğlu B, Nas, Y. (2012). Toplum kaynaklı üriner sistem enfeksiyonu etkeni olarak saptanan Escherichia coli izolatlarının antibiyotik duyarlılıkları. JAREM, 2012; 2(3), 101–103.

2. Gupta K, Hooton TM, Naber KG, et al. International clinical practice guidelines for the treatment of acute uncomplicated cystitis and pyelonephritis in women. Clin Infect Dis. 2011;52(5):e103–e120. doi:10.1093/cid/ciq257

3. Little P, Turner S, Rumsby K, et al. Developing clinical rules to predict urine culture results in primary care. BMJ. 2006;332:143–146. doi:10.1136/bmj.38665.427118.7C

4. Leis JA, Rebick GW, Daneman N, et al. Reducing inappropriate urine cultures: a diagnostic stewardship intervention. JAMA Intern Med. 2014;174(6):876–882. doi:10.1001/jamainternmed.2014.449

5. Morgan DJ, Malani P, Diekema DJ. Diagnostic stewardship—leveraging the laboratory to improve antimicrobial use. JAMA. 2017;318(7):607–608. doi:10.1001/jama.2017.8531

6. Ebell MH, Hansen JG. Diagnosis of urinary tract infections in primary care. BMJ. 2017;359:j4784. doi:10.1136/bmj.j4784

7. Devillé WLJM, Yzermans JC, van Duijn NP, et al. The urine dipstick test useful to rule out infections: a meta-analysis. Fam Pract. 2004;21(4):410–417. doi:10.1093/fampra/cmh412

8. Hurlbut TA, Littenberg B. The diagnostic accuracy of rapid dipstick tests to predict urinary tract infection. Am J Clin Pathol. 1991;96(5):582–588. doi:10.1093/ajcp/96.5.582

9. Stamm WE, Wagner KF, Amsel R, et al. Causes of the acute urethral syndrome in women. N Engl J Med. 1980;303(8):409–415. doi:10.1056/NEJM198008213030803

10. McIsaac WJ, Low DE, Biringer A, et al. The impact of empirical management of acute cystitis on unnecessary antibiotic use. Arch Intern Med. 2002;162(5):600–605. doi:10.1001/archinte.162.5.600

11. Bent S, Nallamothu BK, Simel DL, et al. Does this woman have an acute uncomplicated urinary tract infection? JAMA. 2002;287(20):2701–2710. doi:10.1001/jama.287.20.2701

12. Butler CC, O’Brien K, Pickles T, et al. Point-of-care testing for urinary tract infection in primary care. Lancet Infect Dis. 2021;21(2):199–210. doi:10.1016/S1473-3099(20)30580-0

13. Nicolle LE. Asymptomatic bacteriuria: updated review. Clin Microbiol Rev. 2019;32(2):e00028–18.

14. Drekonja DM, Trautner BW, Amundson C, et al. Effect of a diagnostic stewardship intervention on urine culture ordering. Clin Infect Dis. 2021;72(6):e130–e137. doi:10.1093/cid/ciaa118

15. Bonkat G, Bartoletti R, Bruyère F, et al. EAU guidelines on urological infections 2022. Eur Urol. 2022. doi:10.1016/j.eururo.2022.01.002

16. Wilson ML, Gaido L. Laboratory diagnosis of urinary tract infections in adult patients. Clin Infect Dis. 2004;38(8):1150–1158. doi:10.1086/383029

17. Shrestha NK, Tuohy MJ, Hall GS, et al. Contamination of urine cultures: a persistent problem. J Clin Microbiol. 2021;59(2):e01983–20. doi:10.1128/JCM.01983-20

18. Claeys KC, Zasowski EJ, Trinh TD, et al. Optimal urine culture diagnostic stewardship practice statements: a Delphi consensus. Clin Infect Dis. 2022;75(3):382–390. doi:10.1093/cid/ciac364

19. Kupferwasser DK, Claeys KC, Heil EL, et al. Diagnostic stewardship cutoffs for urinalysis results prior to urine culture ordering. Clin Infect Dis. 2025;80(2):e123–e131. doi:10.1093/cid/ciad694

20. Shrestha NK, Ledeboer NA. Role of pyuria in urinary tract infection diagnosis. Clin Infect Dis. 2020;71(6):e123–e129. doi:10.1093/cid/ciaa050

21. Trautner BW. Management of catheter-associated urinary tract infection. N Engl J Med. 2021;385:2069–2079.

22. Flores-Mireles AL, Walker JN, Caparon M, Hultgren SJ. Urinary tract infections: epidemiology and pathogenesis. Nat Rev Microbiol. 2019;17:269–284.

23. Bossuyt PM, Reitsma JB, Bruns DE, Gatsonis CA, Glasziou PP, Irwig L, et al. The STARD reporting checklist. In: Harwood J, Albury C, Beyer J de, Schlüssel M, Collins G, editors. The EQUATOR network reporting guideline platform [Internet]. The UK EQUATOR Centre; 2025.

